# Advancing Non-Invasive Respiratory Diagnostics: Multiplex Nasal Biomarker Profiling for Stratification of Airway Inflammatory Diseases

**DOI:** 10.1101/2025.08.01.25332809

**Authors:** Tanya Lupancu, Sharmala Thuraisingam, Eldin Rostom, David M. Yen, Brian Wang, Adam M. Damry

## Abstract

This study explores the potential of nasal secretions to serve as a source of biomarkers for diagnosing respiratory conditions. A total of 40 inflammatory biomarkers were detected and quantified in participants with upper respiratory diseases, including chronic rhinosinusitis (CRS), allergic rhinitis, and viral and bacterial infections. The different expression levels of various biomarkers could distinguish CRS participants with and without nasal polyps (i.e. CRP, GzmB, IL-4, MMP-1, MMP-8, SAA and TREM-1), and healthy and rhinitis participants (i.e. CRP, EGF, Eotaxin-1, Fractalkine, IL-1RA, IL-5, IP-10 and TRAIL). CRP, G-CSF, GzmA, IL-1, IL-2, IL-5, IL-8, IL-9, MMP-1, TNFa and TREM-1 protein expression differed between the healthy, viral and bacterial-infected individuals, and EGF, G-CSF, MCP-1, MIP1a and MIP-1b protein expression differed between type 2 and non-type 2 inflammatory cohorts. Significant correlations were also noted between SNOT-22 scores and specific cytokines, such as IP-10 and TRAIL. Despite the heterogeneity of patient diagnoses, these findings highlight the potential of nasal fluid as a readily accessible reflection of respiratory health. Future studies with larger cohorts and standardized methodologies are needed to validate these biomarkers and potentially enable precision diagnosis and improved treatment of respiratory conditions.

## INTRODUCTION

Respiratory diseases are a major global health burden, contributing significantly to morbidity, mortality, and healthcare burden worldwide (1–3). These conditions include a broad spectrum of disorders affecting the upper and lower airway, including acute and chronic infections caused by viral or bacterial pathogens, as well as chronic inflammatory diseases (CRDs) such as chronic rhinosinusitis and asthma. The recent advent of biologics targeting CRDs represents a major advancement in the treatment of these diseases, with a proven impact on population health and quality of life for CRD patients (4, 5). However, despite differing etiology and pathophysiology, many of these highly heterogeneous diseases share overlapping symptoms, such as cough, nasal congestion, wheeze, and mucus overproduction. In addition, current diagnostic practices rely on clinical history, physical examination, imaging, and functional testing though they can lack the sensitivity and specificity required to detect early-stage or mechanistically distinct disease subtypes (6–8). As a result, therapeutic decisions are frequently guided by broad clinical classifications rather than underlying immunopathology, leading to high rates of recalcitrance to treatment and suboptimal use of targeted therapies (9–11).

Chronic rhinosinusitis (CRS) is a heterogenous CRD characterized by persistent inflammation of the nasal and paranasal sinus mucosa, driven by a combination of epithelial barrier dysfunction and immune dysregulation (7, 12, 13). CRS inflammation is predominately a type 2 immune response, defined by eosinophilic infiltration and elevated levels of interleukins IL-4, IL-5, and IL-13 (14, 15). However, additional CRS endotypes exist, involving non-type 2 and low-type 2 inflammation with a predominately neutrophilic-driven profile (16, 17). A similar pattern of immune heterogeneity is observed in asthma, a chronic inflammatory condition of the lower airways, that in many cases, shares the same type 2 cytokine axis and effector cell activity observed in CRS (18, 19). These shared features underscore the concept of the unified airway, which proposes a functional and immunological continuum between the upper and lower respiratory tracts (20–22). The nasal mucosa, as the primary interface with the external environment, plays a sentinel role in detecting and responding to inhaled allergens, pathogens, and pollutants. Nasal fluid, a component of the airway mucus system, traps foreign particles and initiates local immune responses. Rich in cytokines, chemokines, cellular debris, and environmental antigens, it reflects site-specific inflammatory activity (23, 24). The composition of nasal secretions can therefore provide valuable insights into airway immune responses and offers a minimally invasive window into local airway inflammation and infection (25–27).

The COVID-19 pandemic underscored the feasibility of nasal sampling at scale, with widespread use of nasal swabs for diagnostic testing. Furthermore, several studies have demonstrated successful quantification of key type 2 cytokines such as IL-4, IL-5, and IL-13 in nasal samples (28–31), supporting its potential as a matrix for immunological profiling. Given the prevalence of nasal symptoms across many chronic respiratory diseases, nasal fluid is a promising biospecimen for capturing disease-relevant biomarkers and assessing the underlying inflammatory landscape. Therefore, this study aimed to assess whether a broad panel of inflammatory biomarkers can be reliably detected in nasal fluid samples collected from individuals with diverse respiratory diseases, as an initial step towards evaluating the utility of this biospecimen in supporting precision diagnostic approaches.

## MATERIALS AND METHOD

### Study Design and Inclusion/Exclusion Criteria

The study protocol was approved, reviewed and monitored by the Institutional Review Board of Advarra, following the principles of the Helsinki Declaration. Informed consent was obtained from all participants prior to any study-related procedures. Eligible participants were adults aged 18 years or older with moderate rhinorrhea, who were able to blow their nose to secrete 0.5cc of nasal fluid. Participants were excluded from the study if they were unable to blow their nose or could not collect at least 0.5cc of nasal secretions within a 5-minute period, as judged by the investigator.

Participants recruited for the study included those diagnosed by the investigator with the following conditions, based on established diagnostic criteria and clinical evaluation: 1) asthma, according to the Global Initiative for Asthma (GINA), 2) chronic rhinosinusitis (CRS), according to the European Position Paper on Rhinosinusitis and Nasal Polyps (EPOS), 3) rhinitis, 4) type-2 inflammation and 5) respiratory infection.

### Sample and Data Collection

Blown nasal secretions were collected from patients with moderate to severe rhinorrhea. Participants completed a comprehensive patient history and the Sino-Nasal Outcome Test-22 (SNOT-22) questionnaire to assess symptom severity. SNOT-22 consists of 22 questions, each rated from 0 to 5, used to assess the impact of sinonasal symptoms on quality of life. Individual scores are summed to produce a total score ranging from 0 to 110; higher scores indicate greater symptom severity and worse disease-specific quality of life (32). Relevant patient history and demographics are outlined in Table 1.

**Table 1:** Patient Demographics and Clinical Characteristics.

| **Demographic Factor** | **Category** | **Patient (n)** | **Patient (%)** |
| --- | --- | --- | --- |
| **Recruited patients** |  | 58 | 100% |
| **Age Range** | 18-29 | 22 | 38% |
|  | 30-39 | 9 | 16% |
|  | 40-49 | 12 | 21% |
|  | 50-59 | 9 | 16% |
|  | 60+ | 6 | 10% |
| **Sex** | Male | 27 | 47% |
|  | Female | 31 | 53% |
| **Ethnicity** | African American | 3 | 5% |
|  | Asian | 4 | 7% |
|  | Hispanic | 2 | 3% |
|  | White | 39 | 67% |
|  | Multi-racial | 6 | 10% |
|  | Other | 4 | 7% |
| **Asthma Status** | Yes | 12 | 21% |
|  | No | 46 | 79% |
| **Smoking Status** | Current Smoker | 12 | 21% |
|  | Non-Smoker | 46 | 79% |
| **Prior Nasal Surgery** | Yes | 7 | 12% |
|  | No | 51 | 88% |
| **Medication Usage\*** | No Medication | 19 | 33% |
|  | Over the counter | 23 | 40% |
|  | Prescribed for asthma | 9 | 16% |
|  | Prescribed for other | 16 | 28% |
| **Diagnosis** | Healthy | 9 | 16% |
|  | Viral Infection | 25 | 43% |
|  | Bacterial Infection | 6 | 10% |
|  | Chronic Rhinosinusitis with Nasal Polyps | 5 | 9% |
|  | Chronic Rhinosinusitis sans Nasal Polyps | 3 | 5% |
|  | Allergic Rhinitis | 11 | 19% |
|  | Non-Allergic Rhinitis | 5 | 9% |
|  | Type-2 Inflammatory Endotype | 14 | 24% |
|  | Non-Type 2 inflammatory Endotype | 34 | 59% |

\*More information about medication types can be found in Supplementary Table 1.

**Table 2:** Frequency of Detected Immune Mediators in Collected Nasal Secretions.

|  | Total Patients | Healthy Patients | Diseased Patients | LLOD (pg/ml) | ULOD (pg/ml) |
| --- | --- | --- | --- | --- | --- |
| CRP | 95% | 67% | 100% | 7 | 28800 |
| EGF | 88% | 22% | 100% | 2.5 | 10300 |
| Eotaxin-1 | 95% | 67% | 100% | 0.6 | 2850 |
| Eotaxin-2 | 74% | 33% | 82% | 2.2 | 9250 |
| Eotaxin-3 | 78% | 33% | 86% | 0.2 | 900 |
| Fractalkine | 72% | 0% | 86% | 1.1 | 4550 |
| G-CSF | 91% | 56% | 98% | 14.5 | 59500 |
| GM-CSF | 84% | 33% | 94% | 20.6 | 84600 |
| GzmA | 88% | 22% | 100% | 8.4 | 34600 |
| GzmB | 86% | 22% | 98% | 8.1 | 33500 |
| IFNα | 16% | 0% | 18% | 0.6 | 2600 |
| IFNγ | 86% | 44% | 94% | 9.4 | 38800 |
| IL-1RA | 84% | 22% | 96% | 40 | 164100 |
| IL-1α | 83% | 11% | 96% | 0.7 | 3250 |
| IL-2 | 79% | 11% | 92% | 9.8 | 40300 |
| IL-3* | 41% | 0% | 49% | 20.5 | 84000 |
| IL-4 | 91% | 44% | 100% | 15.1 | 62000 |
| IL-5 | 81% | 33% | 90% | 10 | 41200 |
| IL-6 | 81% | 44% | 88% | 12.8 | 52500 |
| IL-8 | 97% | 78% | 100% | 2.5 | 10400 |
| IL-9 | 79% | 11% | 92% | 8.2 | 33700 |
| IL-10 | 86% | 44% | 94% | 1.5 | 6350 |
| IL-12p70 | 95% | 67% | 100% | 7.9 | 32600 |
| IL-13 | 86% | 44% | 94% | 4.1 | 17100 |
| IL-22 | 47% | 0% | 55% | 24.2 | 99200 |
| IL-37 | 91% | 44% | 100% | 3.4 | 14100 |
| IP-10 | 100% | 100% | 100% | 2 | 8500 |
| MCP-1 | 91% | 44% | 100% | 4.2 | 17300 |
| MIP-1α | 76% | 0% | 90% | 1.8 | 7600 |
| MIP-1β | 74% | 0% | 88% | 8 | 33000 |
| MMP-1 | 86% | 56% | 92% | 7 | 28800 |
| MMP-7 | 98% | 89% | 100% | 5.1 | 21200 |
| MMP-8 | 93% | 78% | 96% | 24.8 | 101700 |
| MPO | 100% | 100% | 100% | 76.6 | 313800 |
| SAA | 91% | 44% | 100% | 42.4 | 173800 |
| TNFα | 91% | 44% | 100% | 5.6 | 23200 |
| TNFβ | 17% | 11% | 18% | 6.1 | 25100 |
| TRAIL | 97% | 78% | 100% | 2 | 8250 |
| TREM-1 | 88% | 33% | 98% | 513.2 | 2102300 |
| TREM-2 | 64% | 0% | 76% | 24.7 | 101200 |

### Protein Analysis

Nasal secretion samples were measured using a 40-plex assay, performed by Crux Labs, a contract research organisation based in Australia. The assay was run on the MAGPIX System running xPONENT following manufacturer’s instructions (Thermofisher). Data analysis was carried out using the Belysa Analysis Software (Millpore Sigma).

Analytes measured and their respective lower limit of detection (LLOD) include: C-Reactive protein (CRP; LLOD: 7pg/ml), Epidermal Growth Factor (EGF; LLOD: 2.5pg/ml), Eotaxin-1 (also known as CCL11; LLOD: 0.6pg/ml), Eotaxin-2 (CCL24; LLOD: 2.2pg/ml), Eotaxin-3 (CCL26; LLOD: 0.2pg/ml), Fractalkine (CX3CL1; LLOD: 1.1pg/ml), granulocyte-colony stimulating factor (G-CSF; LLOD: 14.5pg/ml), granulocyte macrophage-colony stimulating factor (GM-CSF; LLOD: 20.6pg/ml), GzmA (Granzyme-A; LLOD: 8.4pg/ml) and GzmB (Granzyme-B; LLOD 8.1pg/ml), interferon (IFN) -alpha (LLOD: 0.6pg/ml), IFNγ (LLOD: 9.4pg/ml), interleukin 1 receptor antagonist (IL-1RA; LLOD: 40pg/ml), interleukin (IL) -2 (LLOD: 9.8pg/ml), IL-3 (LLOD: 20.5pg/ml), IL-4 (LLOD:15.1pg/ml), IL-5 (LLOD:10pg/ml), IL-6 (LLOD: 12.8pg/ml), IL-8 (LLOD: 2.5pg/ml), IL-9 (LLOD: 8.2pg/ml), IL-10 (LLOD: 1.5pg/ml), IL-12p70 (LLOD: 7.9pg/ml), IL-13 (LLOD: 4.1pg/ml), IL-22 (LLOD: 24.2pg/ml), IL-37 (LLOD:3.4pg/ml), interferon gamma-induced protein 10 (IP-10; LLOD: 2pg/ml), monocyte chemoattractant protein-1 (MCP-1, also known as CCL2; LLOD: 4.2pg/ml)), major intrinsic protein-1 (MIP-1) -α (LLOD: 18.5pg/ml), MIP-1β (LLOD: 8pg/ml), matrix metalloproteinases (MMP) -1 (LLOD: 7pg/ml), MMP-7 (LLOD: 5.1pg/ml) , MMP-8 (LLOD: 24.8pg/ml), myeloperoxidase (MPO; LLOD: 76.6pg/ml), serum amyloid A (SAA; LLOD: 42.4pg/ml), tumor necrosis factor (TNF) -α (LLOD: 5.6pg/ml), TNFβ (LLOD: 6.1pg/ml), TNF-Related Apoptosis Inducing Ligand (TRAIL; LLOD: 2pg/ml) and Triggering Receptor Expressed on Myeloid Cells (TREM) -1 (LLOD: 24.7pg/ml) and TREM-2 (LLOD: 24.7pg/ml).

### Data Management and Statistical Analysis

All identifying information was removed and the anonymized data were securely stored in an encrypted, HIPAA-compliant online database (REDCap, Vanderbilt University). To normalize biomarker concentrations, values were scaled using equation 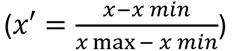. For this purpose, concentrations below the lower limit of detection (LLOD) were imputed with a value equal to LLOD divided by 100. Data analysis was performed using GraphPad Prism 10. Potential relationships between cytokine levels and SNOT-22 scores were investigated using non-parametric Spearman correlations, as variables of interest were not normally distributed. Subgroup analysis was performed using Kruskal-Wallis Tukey’s multiple comparison test to compare the mean concentrations of proteins in patients with allergic and non-allergic rhinitis (AR and NAR, respectively), chronic rhinosinusitis patients with and without nasal polyposis (CRSwNP and CRSsNP, respectively), patients with bacterial versus viral infections, and patients with Type 2 (T2) versus non-T2 inflammation. The alpha level for statistical significance was set at 0.05; with * used to denote p <0.05, ** used to denote p <0.01, *** used to denote p <0.001, **** used to denote p <0.0001.

## RESULTS

### Clinical Characteristics of Study Participants

A total of 58 participants were recruited for this study, with a full breakdown of their clinical characteristics summarised in Table 1. Participant ages ranged from 18 to 70 years, with the mean age and standard deviation being 38.4 and 15.8 years, respectively. There was a near-even distribution of males (47%) and females (53%), and the majority of participants identified as white (67%), with other racial and ethnic groups making up smaller proportions.

Of the 58 participants, 16% were classified as healthy and the remainder had one or more CRD diagnoses, including a respiratory infection (53%), asthma (21%), CRS (14%) or rhinitis (28%). A fifth of participants were current smokers (21%) and two-thirds of participants were taking either over the counter or prescribed medication (67%). Medication usage and types are summarized in Supplementary Table 1.

### Detected Biomarker Concentrations in Collected Nasal Fluid

The concentrations of 40 proteins were measured in all collected nasal fluid samples (Figure 1). Most protein concentrations ranged from 10² to 10⁴ pg/ml, with CRP, IL-1RA, MPO, MMP-8, SAA, and TREM-1 present at concentrations exceeding 10⁴ pg/ml. Notably, CRP, IL-8, MMP-8, and MPO exhibited the widest range of concentrations, spanning from 10⁴ to 10⁸ pg/ml.

**Figure 1:**
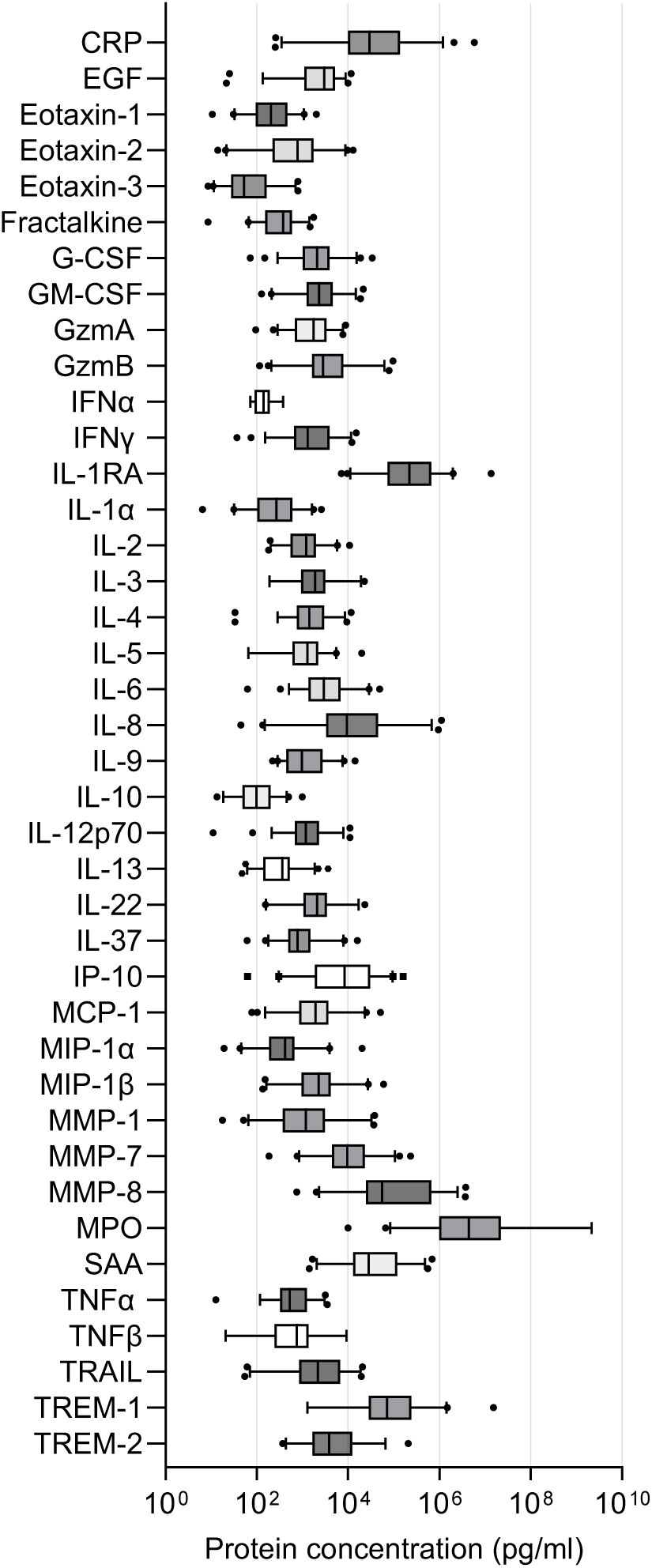
Detected Cytokine Concentrations in Collected Nasal Secretion. The concentration of 40 proteins in the nasal fluid of 58 participants were measured via multiplex immunoassay; the data is presented as boxplot, with 95% confidence intervals (CI). Cytokines measured include C-Reactive protein (CRP), Epidermal Growth Factor (EGF), Eotaxin-1 (also known as CCL11), Eotaxin-2 (CCL24), Eotaxin-3 (CCL26), Fractalkine (CX3CL1), granulocyte-colony stimulating factor (G-CSF), granulocyte macrophage-colony stimulating factor (GM-CSF), granzyme (Gzm) -A and B, interferon (IFN) -α and -γ, interleukin 1 receptor antagonist (IL-1RA), interleukin (IL) -2, -3, -4, -5, -6, -8, -9, -10, -12p70, -13, -22, -37, interferon gamma-induced protein 10 (IP-10), monocyte chemoattractant protein-1 (MCP-1, also known as CCL2), major intrinsic protein-1 (MIP-1) - α, -β, matrix metalloproteinases (MMP) -1, -7, -8, myeloperoxidase (MPO), serum amyloid A (SAA), tumor necrosis factor (TNF) - α, -β, TNF-Related Apoptosis Inducing Ligand (TRAIL) and Triggering Receptor Expressed on Myeloid Cells (TREM) -1 and -2.

Not all 40 proteins were detected in all 58 nasal secretion samples. The majority of the proteins in the panel were inflammatory cytokines and only those detected in more than 80% of samples from patients with CRDs were included in subsequent analyses. A total of 36 proteins met this criterion, with IFNα, IL-22, TNFβ, and TREM-2 being excluded at this stage. Amongst the 36 proteins, Fractalkine, IL-3, MIP-1α, and MIP-1β were only detected in samples from patients with CRDs, while the remaining 33 biomarkers were also detected in healthy patients. However, no biomarkers were observed at a higher prevalence in samples from healthy patients relative to those from patients with CRDs.

### Correlation of SNOT-22 scores and Biomarker Concentrations

In addition to nasal secretion collection, participants were asked to complete the SNOT-22 questionnaire. Just over half the study cohort (n=31, 53%) completed the SNOT-22 questionnaire; the total mean score was 39 (95% CI: 30.7 to 47.7), the mean SNOT-22 score for diseased participants was 44.8 (95% CI:36.1 to 53.6, n=25), and the mean SNOT-22 score for healthy participants was and 15.6 (95% CI: 0.64 to 30.69, n=6) (Supplementary Figure 1). Statistically significant correlations were observed between SNOT-22 scores and concentrations of Eotaxin-2 , Fractalkine, IP-10 and TRAIL (Table 3). Eotaxin-2 (r= -0.52; 95% CI: -0.78 to -0.11; p value= 0.01) and Fractalkine (r= -0.6025; 95% CI: -0.8136 to - 0.2513; p value= 0.002) were negatively correlated to SNOT-22 scores, while IP-10 (r=0.45; 95% CI 0.01 to 0.70; p= 0.01) and TRAIL (r= 0.48; 95% CI= 0.13 to 0.72; p=0.01) were positively correlated to SNOT-22 scores.

**Table 3:** Correlation between SNOT-22 Scores and Cytokine Concentrations.

| **Cytokine** | **Spearman Correlation**<br>**R Value** | **95% CI** | **P VALUE** | **XY Pairs** |
| --- | --- | --- | --- | --- |
| CRP | 0.10 | -0.30 to 0.47 | 0.60 | 28 |
| EGF | -0.30 | -0.61 to 0.11 | 0.13 | 27 |
| Eotaxin-1 | 0.03 | -0.35 to 0.40 | 0.88 | 29 |
| Eotaxin-2 | -0.52 | -0.78 to -0.11 | 0.01 | 22 |
| Eotaxin-3 | -0.27 | -0.62 to 0.15 | 0.19 | 24 |
| Fractalkine | -0.60 | -0.82 to -0.25 | 0.002 | 24 |
| G-CSF | -0.04 | -0.42 to 0.36 | 0.86 | 27 |
| GM-CSF | -0.14 | -0.51 to 0.27 | 0.49 | 26 |
| GzmA | 0.06 | -0.35 to 0.45 | 0.77 | 26 |
| GzmB | -0.18 | -0.55 to 0.24 | 0.38 | 25 |
| IFNγ | -0.21 | -0.56 to 0.20 | 0.29 | 26 |
| IL-1Rα | -0.08 | -0.45 to 0.32 | 0.71 | 27 |
| IL-1α | -0.26 | -0.60 to 0.15 | 0.20 | 26 |
| IL-2 | -0.31 | -0.63 to 0.11 | 0.14 | 25 |
| IL-3 | -0.44 | -0.79 to 0.14 | 0.12 | 14 |
| IL-4 | -0.20 | -0.55 to 0.21 | 0.31 | 27 |
| IL-5 | -0.19 | -0.56 to 0.24 | 0.38 | 24 |
| IL-6 | -0.30 | -0.63 to 0.13 | 0.16 | 24 |
| IL-8 | 0.07 | -0.31 to 0.42 | 0.72 | 31 |
| IL-9 | -0.37 | -0.67 to 0.048 | 0.07 | 25 |
| IL-10 | -0.17 | -0.53 to 0.25 | 0.42 | 26 |
| IL-12p70 | -0.22 | -0.56 to 0.19 | 0.27 | 27 |
| IL-13 | -0.34 | -0.65 to 0.07 | 0.09 | 26 |
| IL-37 | -0.16 | -0.52 to 0.25 | 0.43 | 27 |
| IP-10 | 0.45 | 0.10 to 0.70 | 0.01 | 31 |
| MCP-1 | -0.07 | -0.45 to 0.33 | 0.71 | 27 |
| MIP-1α | -0.24 | -0.60 to 0.19 | 0.26 | 24 |
| MIP-1β | -0.34 | -0.66 to 0.09 | 0.11 | 24 |
| MMP-1 | -0.04 | -0.43 to 0.37 | 0.86 | 26 |
| MMP-7 | -0.09 | -0.45 to 0.29 | 0.63 | 30 |
| MMP-8 | 0.06 | -0.32 to 0.43 | 0.75 | 29 |
| MPO | 0.03 | -0.34 to 0.39 | 0.87 | 31 |
| SAA | 0.13 | -0.27 to 0.50 | 0.51 | 27 |
| TNFα | -0.06 | -0.44 to 0.34 | 0.75 | 27 |
| TRAIL | 0.48 | 0.13 to 0.72 | 0.01 | 30 |
| TREM-1 | -0.18 | -0.53 to 0.23 | 0.37 | 27 |

### Association of Biomarker Concentrations to CRD Classifications

With SNOT-22 scores being a subjective and poorly quantitative assessment of nasal health, biomarker concentrations were correlated to CRD diagnoses across subsets of patients. In the case of rhinitis, a heatmap was generated to represent normalized protein concentrations and assess whether these markers could stratify participants into healthy, allergic rhinitis (AR), or non-allergic rhinitis (NAR) groups (Figure 2). Biomarker levels were variably upregulated in the AR and NAR rhinitis cohorts, and the mean protein expression of CRP, EGF, Eotaxin-1, Fractalkine, IL-1RA, IL-5, IP-10 and TRAIL differed between participants in the healthy and rhinitis cohorts (see Supplementary Table 2). However, no single biomarker could reliably stratify participants into the three groups.

**Figure 2:**
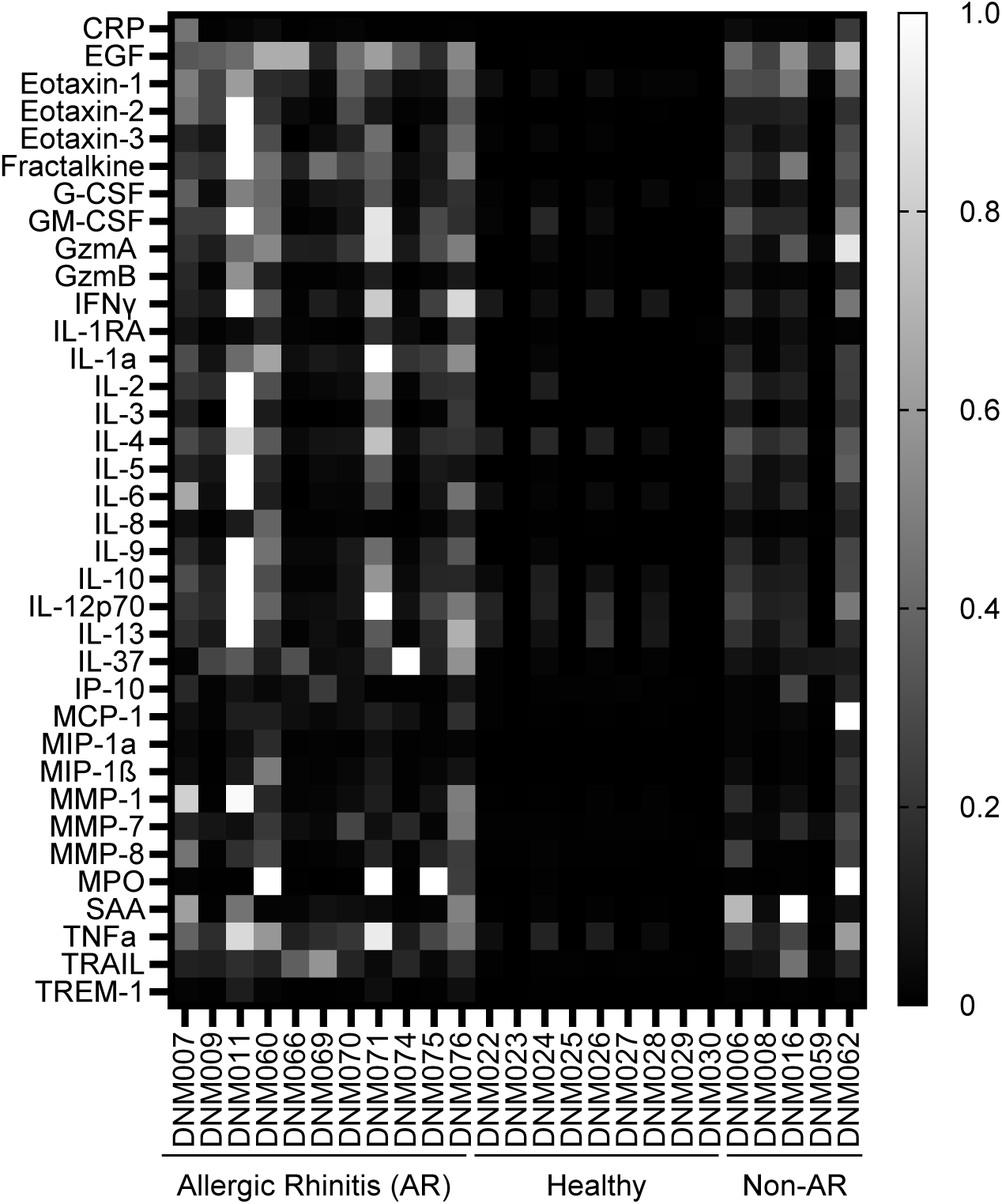
Heatmap of Cytokine Concentrations across Inflammatory Rhinitis Patient Cohort. Normalised cytokine concentrations were visualized using a heatmap. Patient cohorts plotted included those with allergic rhinitis (AR) (n=11), healthy patients (n=9) and patients with non-allergic rhinitis (NAR) (n=5).

Next, a similar analysis was performed to identify biomarkers that could distinguish types of chronic rhinosinusitis. Another heatmap was used to stratify participants into healthy, chronic rhinosinusitis sans nasal polyps (CRSsNP) and chronic rhinosinusitis with nasal polyps (CRSwNP) cohorts based on their normalized protein concentrations (Figure 3).

**Figure 3:**
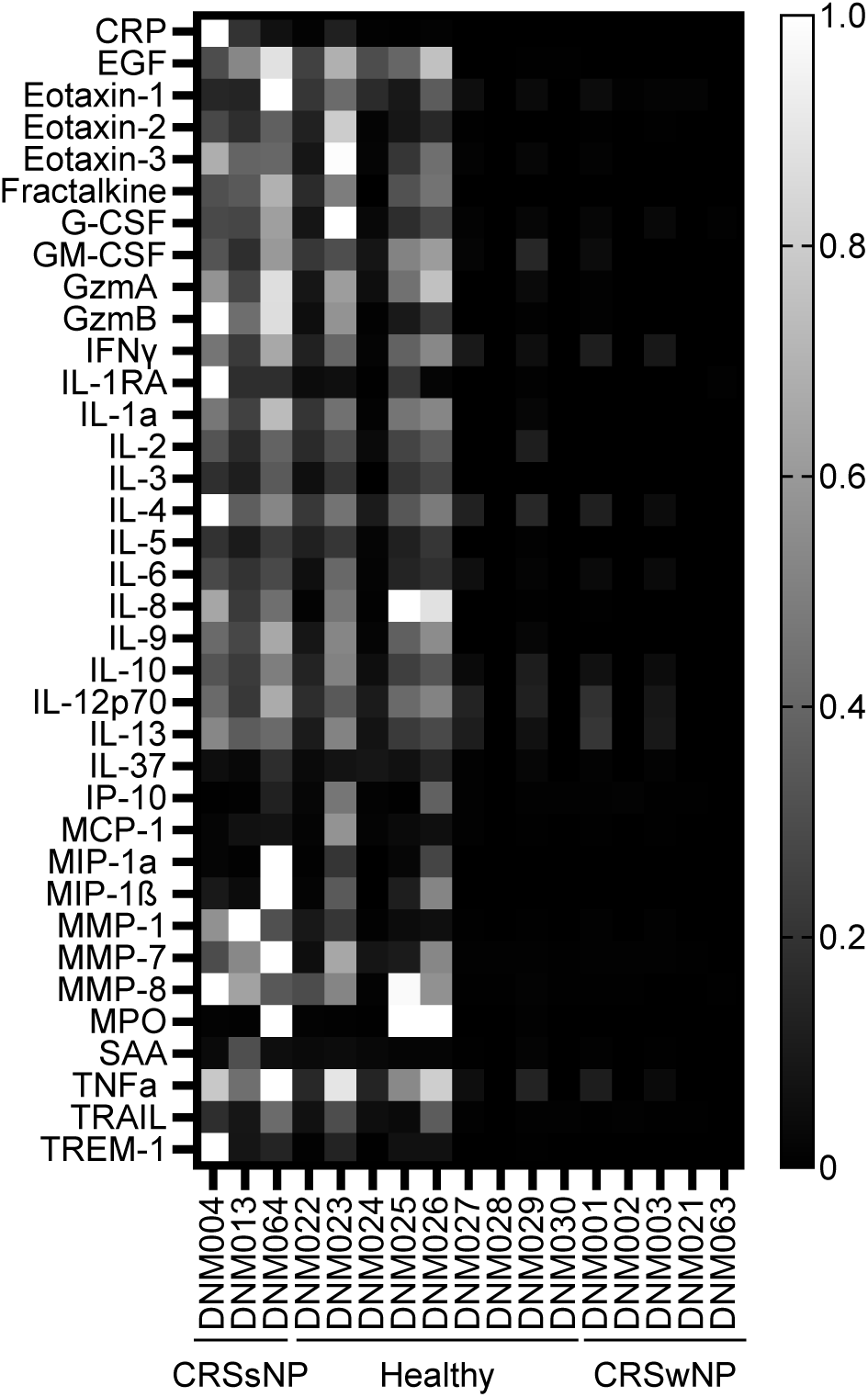
Heatmap of Cytokine Concentrations across Chronic Rhinosinusitis (CRS) Patient Cohort. Normalised cytokine concentrations were visualized using a heatmap. Patient cohorts plotted included those with chronic rhinosinusitis sans nasal polyps (CRSsNP) (n=3), healthy patients (n=9) and patients with chronic rhinosinusitis with nasal polyps (CRSwNP) (n=5).

Compared to the healthy group, biomarker concentrations were elevated in both CRS cohorts. Statistical analysis showed that the concentrations of select proteins (i.e. CRP, GzmB, IL-4, MMP-1, MMP-8, SAA and TREM-1) were unique to specific cohorts (Supplementary Table 3); however, the CRS cohort was the smallest in the study, which limits interpretation.

Inflammatory biomarkers are also known to be upregulated in response to pathogenic infection (33, 34). Biomarker concentrations were next analyzed to verify if they could be used to stratify patients into healthy, viral or bacterial infection cohorts (Figure 4). Significant differences in the protein expression of CRP, G-CSF, GzmA, IL-1, IL-2, IL-5, IL-8, IL-9, MMP-1, TNFa and TREM-1 were observed between the 3 cohorts (Supplementary Table 4).

**Figure 4:**
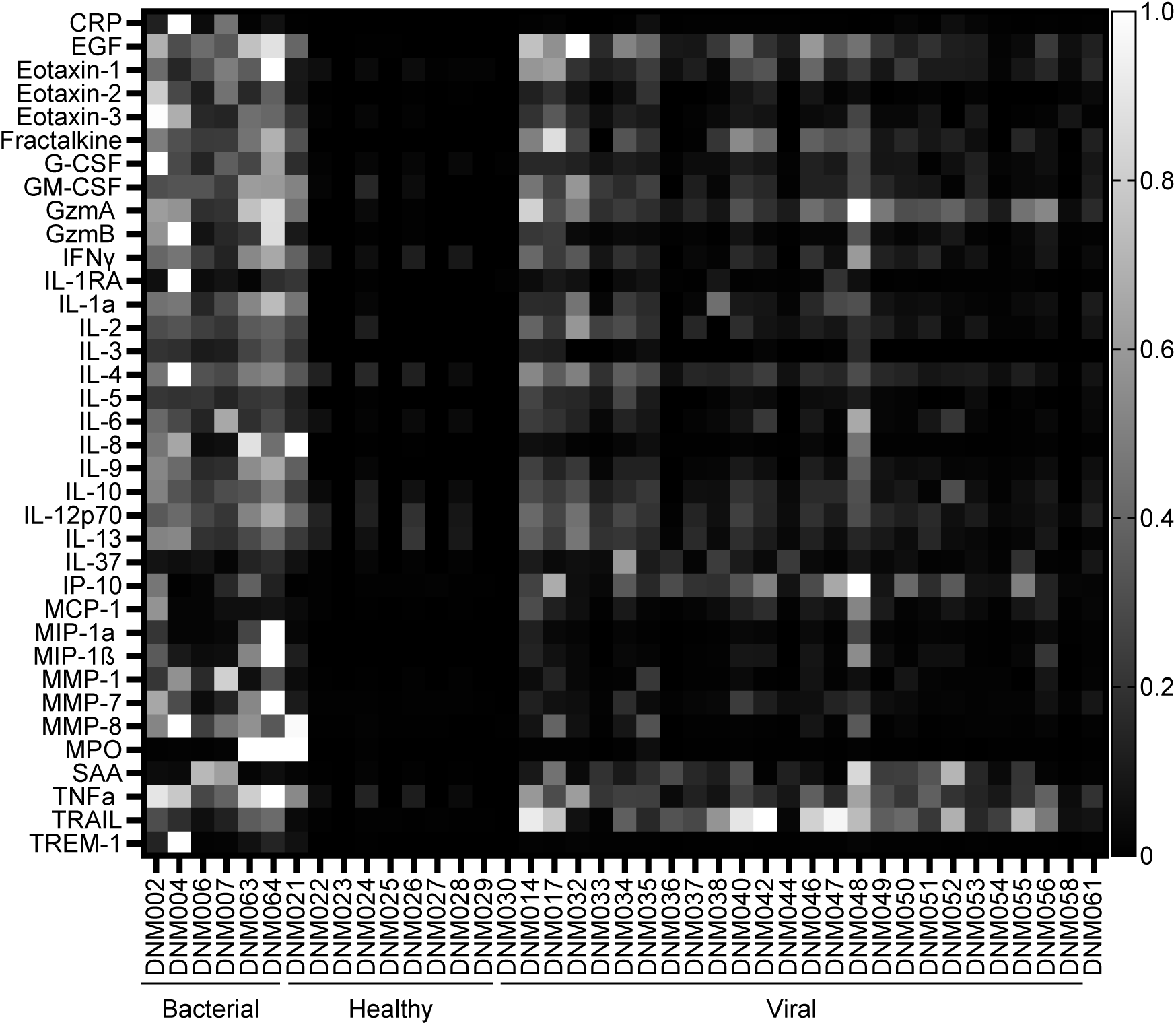
Heatmap of Cytokine Concentrations Across Infectious Rhinosinusitis Patient Cohort. Normalised cytokine concentrations were visualized using a heatmap. Patient cohorts plotted included those with bacterial infection (n=6), healthy patients (n=9) and patients with viral infection (n=25).

Finally, the normalized protein concentration of participants with T2 and non-T2 inflammatory endotypes were visualised in a heatmap to verify whether these biomarkers can be used to stratify T2 from non-T2 CRDs (Figure 5). Higher levels of protein expression were generally seen in the T2 cohort, with significant differences in EGF (p=0.03), MCP-1 (0.02), MIP1a (p=0.02) and MIP-1b (p=0.03) observed between the two cohorts (Supplementary Figure 5).

**Figure 5:**
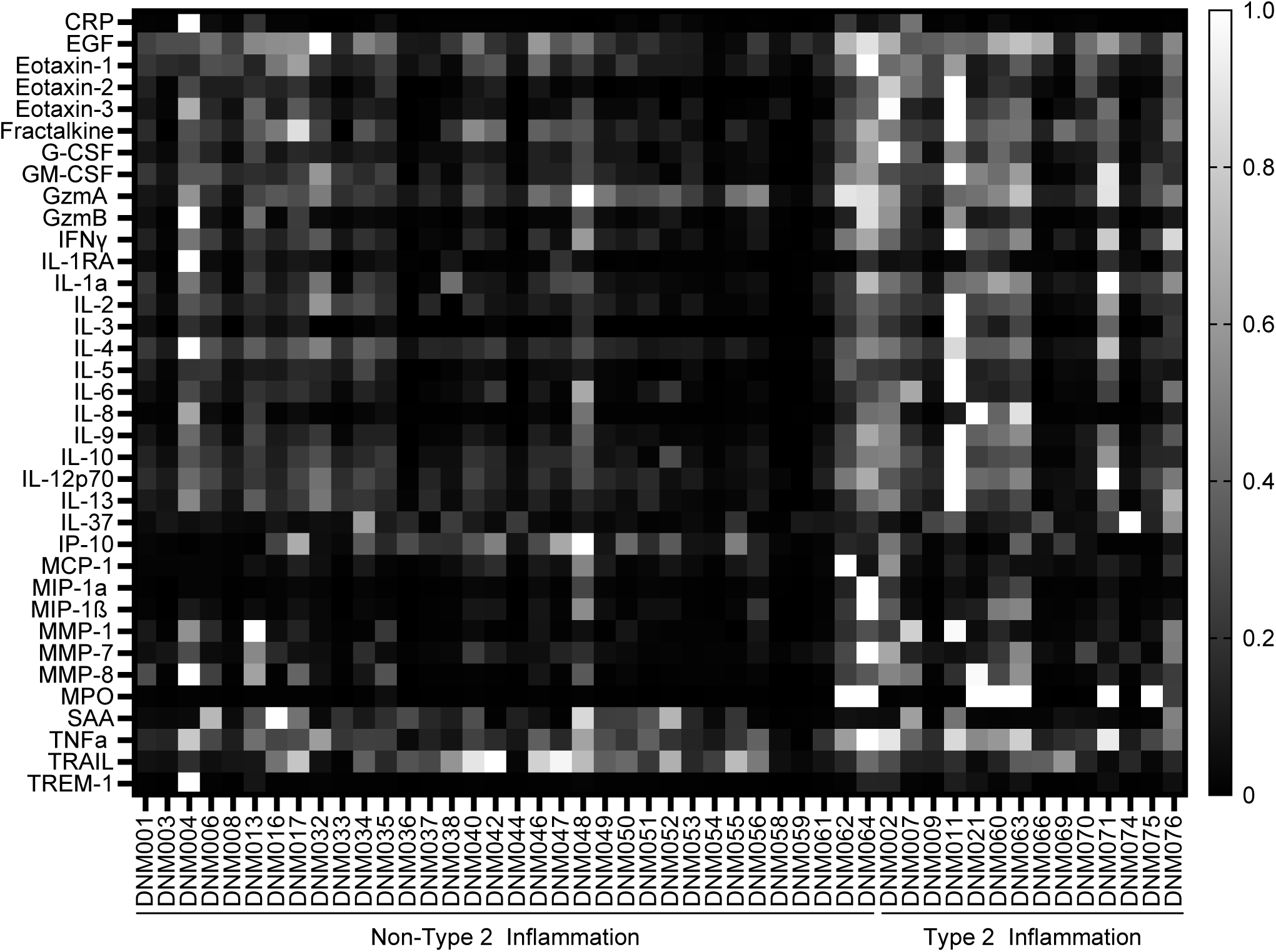
Heatmap of Cytokine Concentrations Across Th-2 mediated Inflammatory Patient Cohort. Normalised cytokine concentrations were visualized using a heatmap. Patient cohorts plotted included those with non-Type 2 inflammation (n=32) and Type 2 Inflammation (n=14).

## DISCUSSION

Nasal fluid is a rich source of biomarkers and in the present study, 40 inflammatory protein biomarkers were detected in samples collected from patients with varying CRDs. While the patient cohorts for each disease group were generally too small to make clear correlative associations for single biomarkers, distinct biomarker trends were observed between healthy and diseased patients.

In this work, we investigated whether quantitative biomarker detection could recapitulate scores from the qualitative and imprecise SNOT-22 questionnaire. SNOT-22 is used by clinicians to assess the severity of symptoms and quality of life of patients with nasal and sinus-related conditions, with significant changes also used to predict sinus surgery outcomes and the efficacy of treatments (32, 35, 36). Previous studies have demonstrated correlation between cytokine concentrations and SNOT-22 scores, with elevated protein concentrations of IL-5 and IL-13 found to positively correlate to SNOT-22 scores in CRS patients (37, 38). Our study, however, included only eight CRS patients, making it unsurprising that we did not find a statistically significant correlation between SNOT-22 scores and IL-5 or IL-13. On the other hand, we did observe negative correlations with eotaxin-2 and fractalkine, and positive correlations with IP-10 and TRAIL, despite the heterogeneity of patient diagnoses. These results highlight that SNOT-22 scores may offer insight into biological changes, while similarly demonstrating that a panel of selected biomarkers could complement or, in the long term, potentially replace the SNOT-22 questionnaire in assessing these changes more accurately and less subjectively.

As CRDs are highly heterogenous diseases, it is also important to identify causes and endotypes to administer effective treatments. To do so, several studies have sought to stratify CRS patients into clusters based on cytokine protein levels in sinonasal surgical tissue. One study identified ten different clusters in 173 CRS patients using IL-5, ECP, IgE and albumin (39), while another identified three clusters in 26 patients based on their expression of Th1/Th17 cytokines, Th2 cytokines and chemokines/ growth factors (40). In this work, we have demonstrated that a diverse range of inflammatory biomarkers can be detected in nasal fluid, including many that present statistical differences between various clusters. Our results therefore show that a similar stratification to surgical sample-based studies may be possible with less invasive means by quantifying biomarkers in easily-accessible blown nasal fluid instead. Indeed, while no single biomarker could perfectly account for differences between clusters, the trends observed on a relatively small panel of biomarkers highlights that multi-marker correlations discovered through larger panels may be successful in achieving sensitive and specific stratification of CRDs.

Of note amongst tested clusters, the lack of definitive diagnostic tests to differentiate viral from bacterial respiratory infections remains a significant challenge in healthcare (41, 42). Incorrect diagnoses often result in delayed or incorrect treatment, including the overuse of antibiotics (43), and while PCR and culturing remain the gold standards for diagnosis, confirmation can take days (44). To address these needs, several projects have sought to identify protein-based biomarkers and host-derived gene expression signatures to distinguish between viral and bacterial infections. These studies are complicated by the heterogeneous nature of these infections, with different pathogens presenting different biomarker patterns (45, 46).

Nonetheless, several blood-based infection biomarkers have been identified with varying sensitivity and specificity, with serum IP-10 and TRAIL having been found to be associated with high viral loads (47–50). Our study highlights similar trends in nasal fluid, with both biomarkers being on average elevated in the nasal secretions of our virally infected participants. Similarly, markers like TNFα and MPO have shown trends of elevation in subsets of bacterially infected patients’ nasal fluid (51), suggesting once again that a combined biomarker sensing approach that contrasts biomarkers that are differentially elevated in viral and bacterial infections may achieve the necessary sensitivity and specificity for rapid clinical differentiation of these infections.

Overall, this study demonstrated the potential of nasal secretions as a valuable source of biomarkers. While blood has traditionally been the preferred medium for biomarker research, supported by an extensive body of literature, nasal secretions offer distinct advantages for CRD diagnosis and monitoring. Sourced directly from the respiratory system, nasal fluid could provide a more localised and direct reflection of respiratory health, potentially capturing nuanced physiological changes that systemically-representative blood samples may miss. Additionally, the easy and non-invasive collection of nasal fluid makes it particularly suited for frequent testing and large-scale studies, although standardized collection methods need to be established to make such studies reliable and reproducible (52). As research on nasal fluid advances, it stands to be a critical tool for identifying new biomarkers and improving the diagnosis and treatment of respiratory conditions that remain challenging to detect through traditional testing.

## LIST OF ABBREVIATIONS

ANOVA: Analysis of Variance
CCL11: C-C Motif Chemokine Ligand 11
CCL24: C-C Motif Chemokine Ligand 24
CCL26: C-C Motif Chemokine Ligand 26
COVID-19: Coronavirus Disease 2019
CRD: Chronic Respiratory Disease
CRS: Chronic Rhinosinusitis
CX3CL1: C-X3-C Motif Chemokine Ligand 1
EGF: Epidermal Growth Factor
G-CSF: Granulocyte Colony-Stimulating Factor
GM-CSF: Granulocyte Macrophage Colony-Stimulating Factor
GzmA: Granzyme A
GzmB: Granzyme B
IFN-α: Interferon-Alpha
IFNγ: Interferon-Gamma
IL-1RA: Interleukin-1 Receptor Antagonist
IL-2: Interleukin-2
IL-3: Interleukin-3
IL-4: Interleukin-4
IL-5: Interleukin-5
IL-6: Interleukin-6
IL-8: Interleukin-8
IL-9: Interleukin-9
IL-10: Interleukin-10
IL-12p70: Interleukin-12p70
IL-13: Interleukin-13
IL-22: Interleukin-22
IL-37: Interleukin-37
IP-10: Interferon Gamma-Induced Protein 10
MCP-1: Monocyte Chemoattractant Protein 1
MIP-1α: Macrophage Inflammatory Protein 1-Alpha
MIP-1β: Macrophage Inflammatory Protein 1-Beta
MMP-1: Matrix Metalloproteinase 1
MMP-7: Matrix Metalloproteinase 7
MMP-8: Matrix Metalloproteinase 8
MPO: Myeloperoxidase
SAA: Serum Amyloid A
SNOT-22: Sino-Nasal Outcome Test
SPA: Specialty Physician Associates
TNF-α: Tumor Necrosis Factor Alpha
TNFβ: Tumor Necrosis Factor Beta
TRAIL: TNF-Related Apoptosis Inducing Ligand
TREM-1: Triggering Receptor Expressed on Myeloid Cells 1
TREM-2: Triggering Receptor Expressed on Myeloid Cells 2

## DECLARATIONS

### Ethics approval and consent to participate

Informed consent was obtained from patients.

### Consent for publication

Not applicable

### Availability of data and materials

All data generated or analysed during this study are included in this published article and its supplementary information files.

### Competing Interests

All authors receive financial compensation from Diag-Nose Medical, which funded this research. As such, they may have a financial or professional interest in the results. This relationship is disclosed in the author affiliations. No additional competing interests are declared.

### Funding

This work was funded by Diag-nose Medical.

### Authors’ contributions

TL conducted the formal analysis and contributed to conceptualisation, drafting, review and editing of the manuscript. ST contributed to the formal analysis and editing of the manuscript. AD contributed to the conceptualization and design of the study, and participated in manuscript review and editing. ER, DY, and BW contributed to the conceptualization and study design. All authors read and approved the final manuscript.

## Acknowledgements

We thank L.Kaur and A.Yusim of Crux Biolabs for performing the experiments.

## Notes

### Author Declarations

Ethics IRB of Advarra gave ethical approval for this work.

