## Supplemental Data for "Advancing Non-Invasive Respiratory Diagnostics: Multiplex Nasal Biomarker Profiling for Stratification of Airway Inflammatory Diseases"


**Author Full Names**

Tanya Lupancu[1], Sharmala Thuraisingam[1,2], Eldin Rostom[1], David M. Yen[1,3], Brian Wang[1,4], Adam M. Damry[1,5].

**Affiliations.**

1. Diag-Nose Medical, Notting Hill, 3168, Victoria, Australia.
2. Department of Surgery, The University of Melbourne, Fitzroy, 3065, Victoria, Australia.
3. Specialty Physician Associates, Bethlehem, 18107, Pennsylvania, United States
4. Houston Methodist Hospital, Department of Otolaryngology – Head and Neck Surgery, Houston, TX, United States
5. Department of Chemistry, University of Ottawa, Ottawa, ON K1N 6N5, Canada

**Corresponding author**

Adam M. Damry;

**Supplementary Table 1: Medications prescribed for study participants**

| Prescribed For Asthma | Over The Counter | Prescribed For Non-Respiratory Indications |
|---|---|---|
| **Antibiotics:** amoxicillin, azithromycin, sulfonamides | **Antihistamines*** | **Beta blocker medication:** atenolol, metoprolol, propranolol |
| **Anticholinergic/ bronchodilator:** ipratropium, salbutamol, tiotropium bromide monohydrate | **Analgesic**: Tylenol | **CNS-targeting medicine:** muscle relaxants (cyclobenzaprine), stimulant (amphetamine) sedatives (zolpidem and alprazolam), pain management (pregabalin and tramadol), antidepressants (duloxetine, trazodone, venlafaxine, Lexapro, Zoloft, Adderall |
| **Leukotriene modifier:** montelukast | **Decongestants**: Cold and flu medicine (oroidin, NyQuil, Sudafed) | **Diuretics:** hydrochlorothiazide and Olmesartan/ triamterene |
| **Inhaled Cortico/Steroids:** Budesonide, formoterol, fluticasone Propionate | **NSAIDS:** aspirin, celecoxib, ibuprofen | **Gastric reflux medication:** esomeprazole, famotidine, lansoprazole, omeprazole, pantoprazole |
| **Biologics:** dupilumab | | **High blood pressure medication:** amlodipine, atenolol, enalapril, fosinopril, lisinopril, losartan, olmesartan, rosuvastatin, telmisartan, valsartan |
| | | **Migraine medication:** fremanezumab, rizatriptan, sumatriptan, topiramate, triptan |
| | | **Statins:** atorvastatin, rosuvastatin, simvastatin |
| | | **Type 2 diabetes medication:** canagliflozin, metformin, semaglutide |
| | | **Other**: adapalene and clobetasol creams, conjugated estrogens, finasteride, hypothyroidism medication (levothyroxine), oral contraception, supplements**, tamsulosin |

*Afrin, Allegra, Azelastine, Benadryl, Clarinex, Claritin, Patanol, Zicam, Zyrtec

**Biotin, calcium, antioxidant, cranberry, fish oil, glucosamine, probiotic, magnesium, multivitamin, melatonin

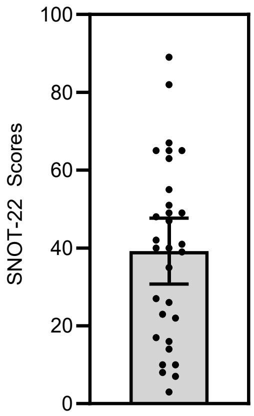


**Supplementary Figure 1: Range of SNOT-22 scores collected**

The data is presented as boxplot, with 95% CI, n=31.

| Supplementary Table 2: Kruskal-Wallis test comparing inflammatory rhinitis cohorts, with Dunn's pairwise comparison. Cytokine | Comparison Cohort | Adjusted P Value |
|---|---|---|
| **CRP** | AR vs. Healthy | 0.001 |
| | AR vs. NAR | >0.999 |
| | Healthy vs. NAR | 0.006 |
| **EGF** | AR vs. Healthy | <0.001 |
| | AR vs. NAR | >0.999 |
| | Healthy vs. NAR | 0.005 |
| **Eotaxin-1** | AR vs. Healthy | 0.003 |
| | AR vs. NAR | >0.999 |
| | Healthy vs. NAR | 0.018 |
| **Eotaxin-2** | AR vs. Healthy | <0.001 |
| | AR vs. NAR | >0.999 |
| | Healthy vs. NAR | 0.073 |
| **Eotaxin-3** | AR vs. Healthy | <0.001 |
| | AR vs. NAR | >0.999 |
| | Healthy vs. NAR | 0.073 |
| **Fractalkine** | AR vs. Healthy | <0.001 |
| | AR vs. NAR | >0.999 |
| | Healthy vs. NAR | 0.041 |
| **G-CSF** | AR vs. Healthy | 0.002 |
| | AR vs. NAR | >0.999 |
| | Healthy vs. NAR | 0.158 |
| **GM-CSF** | AR vs. Healthy | 0.004 |
| | AR vs. NAR | >0.999 |
| | Healthy vs. NAR | 0.068 |
| **GzmA** | AR vs. Healthy | <0.001 |
| | AR vs. NAR | >0.999 |
| | Healthy vs. NAR | 0.019 |
| **GzmB** | AR vs. Healthy | 0.001 |
| | AR vs. NAR | >0.999 |

| | | |
|---|---|---|
| | Healthy vs. NAR | 0.060 |
| **IFNγ** | AR vs. Healthy | 0.018 |
| | AR vs. NAR | >0.999 |
| | Healthy vs. NAR | 0.281 |
| **IL-1RA** | AR vs. Healthy | <0.001 |
| | AR vs. NAR | >0.999 |
| | Healthy vs. NAR | 0.048 |
| **IL-1a** | AR vs. Healthy | <0.001 |
| | AR vs. NAR | 0.626 |
| | Healthy vs. NAR | 0.089 |
| **IL-2** | AR vs. Healthy | 0.001 |
| | AR vs. NAR | >0.999 |
| | Healthy vs. NAR | 0.054 |
| **IL-3** | AR vs. Healthy | 0.016 |
| | AR vs. NAR | >0.999 |
| | Healthy vs. NAR | 0.163 |
| **IL-4** | AR vs. Healthy | 0.012 |
| | AR vs. NAR | >0.999 |
| | Healthy vs. NAR | 0.053 |
| **IL-5** | AR vs. Healthy | 0.001 |
| | AR vs. NAR | >0.999 |
| | Healthy vs. NAR | 0.017 |
| **IL-6** | AR vs. Healthy | 0.055 |
| | AR vs. NAR | >0.999 |
| | Healthy vs. NAR | 0.156 |
| **IL-8** | AR vs. Healthy | 0.001 |
| | AR vs. NAR | >0.999 |
| | Healthy vs. NAR | 0.016 |
| **IL-9** | AR vs. Healthy | <0.001 |
| | AR vs. NAR | >0.999 |
| | Healthy vs. NAR | 0.052 |
| **IL-10** | AR vs. Healthy | 0.011 |
| | AR vs. NAR | >0.999 |
| | Healthy vs. NAR | 0.185 |
| **IL-12p70** | AR vs. Healthy | 0.032 |
| | AR vs. NAR | >0.999 |
| | Healthy vs. NAR | 0.377 |

| | | |
|---|---|---|
| **IL13** | AR vs. Healthy<br>AR vs. NAR<br>Healthy vs. NAR | 0.086<br>>0.999<br>0.514 |
| **IL-37** | AR vs. Healthy<br>AR vs. NAR<br>Healthy vs. NAR | <0.001<br>0.890<br>0.061 |
| **IP-10** | AR vs. Healthy<br>AR vs. NAR<br>Healthy vs. NAR | 0.005<br>>0.999<br>0.025 |
| **MCP-1** | AR vs. Healthy<br>AR vs. NAR<br>Healthy vs. NAR | 0.001<br>0.833<br>0.091 |
| **MIP-1a** | AR vs. Healthy<br>AR vs. NAR<br>Healthy vs. NAR | <0.001<br>>0.999<br>0.031 |
| **MIP-1ß** | AR vs. Healthy<br>AR vs. NAR<br>Healthy vs. NAR | <0.001<br>>0.999<br>0.054 |
| **MMP-1** | AR vs. Healthy<br>AR vs. NAR<br>Healthy vs. NAR | <0.001<br>>0.999<br>0.046 |
| **MMP-7** | AR vs. Healthy<br>AR vs. NAR<br>Healthy vs. NAR | <0.001<br>>0.999<br>0.014 |
| **MMP-8** | AR vs. Healthy<br>AR vs. NAR<br>Healthy vs. NAR | 0.002<br>0.949<br>0.218 |
| **MPO** | AR vs. Healthy<br>AR vs. NAR<br>Healthy vs. NAR | 0.006<br>>0.999<br>0.055 |
| **SAA** | AR vs. Healthy<br>AR vs. NAR<br>Healthy vs. NAR | 0.006<br>>0.999<br>0.055 |
| **TNFa** | AR vs. Healthy<br>AR vs. NAR<br>Healthy vs. NAR | <0.001<br>>0.999<br>0.068 |
| **TRAIL** | AR vs. Healthy | <0.001 |

| | | |
|---|---|---|
| | AR vs. NAR | >0.999 |
| | Healthy vs. NAR | 0.019 |
| **TREM-1** | AR vs. Healthy | <0.001 |
| | AR vs. NAR | >0.999 |
| | Healthy vs. NAR | 0.092 |

**Supplementary Table 3: Kruskal-Wallis test comparing CRS cohorts, with Dunn's pairwise comparisons .**

| Cytokine | Comparison Cohort | Adjusted P Value |
|---|---|---|
| **CRP** | CRSsNP vs. Healthy | 0.096 |
| | CRSsNP vs. CRSwNP | 0.005 |
| | Healthy vs. CRSwNP | 0.372 |
| **EGF** | CRSsNP vs. Healthy | 0.693 |
| | CRSsNP vs. CRSwNP | 0.015 |
| | Healthy vs. CRSwNP | 0.073 |
| **Eotaxin-1** | CRSsNP vs. Healthy | 0.869 |
| | CRSsNP vs. CRSwNP | 0.071 |
| | Healthy vs. CRSwNP | 0.267 |
| **Eotaxin-2** | CRSsNP vs. Healthy | 0.205 |
| | CRSsNP vs. CRSwNP | 0.023 |
| | Healthy vs. CRSwNP | 0.573 |
| **Eotaxin-3** | CRSsNP vs. Healthy | 0.515 |
| | CRSsNP vs. CRSwNP | 0.016 |
| | Healthy vs. CRSwNP | 0.134 |
| **Fractalkine** | CRSsNP vs. Healthy | 0.257 |
| | CRSsNP vs. CRSwNP | 0.022 |
| | Healthy vs. CRSwNP | 0.435 |
| **G-CSF** | CRSsNP vs. Healthy | 0.261 |
| | CRSsNP vs. CRSwNP | 0.028 |
| | Healthy vs. CRSwNP | 0.519 |
| **GM-CSF** | CRSsNP vs. Healthy | 0.797 |
| | CRSsNP vs. CRSwNP | 0.031 |
| | Healthy vs. CRSwNP | 0.126 |
| **GzmA** | CRSsNP vs. Healthy | 0.424 |
| | CRSsNP vs. CRSwNP | 0.022 |
| | Healthy vs. CRSwNP | 0.241 |
| **GzmB** | CRSsNP vs. Healthy | 0.125 |
| | CRSsNP vs. CRSwNP | 0.007 |
| | Healthy vs. CRSwNP | 0.378 |
| **IFNγ** | CRSsNP vs. Healthy | 0.284 |
| | CRSsNP vs. CRSwNP | 0.033 |
| | Healthy vs. CRSwNP | 0.547 |
| **IL-1RA** | CRSsNP vs. Healthy | 0.166 |

| | | |
|---|---|---|
| | CRSsNP vs. CRSwNP | 0.011 |
| | Healthy vs. CRSwNP | 0.391 |
| **IL-1a** | CRSsNP vs. Healthy | 0.339 |
| | CRSsNP vs. CRSwNP | 0.011 |
| | Healthy vs. CRSwNP | 0.166 |
| **IL-2** | CRSsNP vs. Healthy | 0.339 |
| | CRSsNP vs. CRSwNP | 0.011 |
| | Healthy vs. CRSwNP | 0.166 |
| **IL-3** | CRSsNP vs. Healthy | 0.349 |
| | CRSsNP vs. CRSwNP | 0.030 |
| | Healthy vs. CRSwNP | 0.400 |
| **IL-4** | CRSsNP vs. Healthy | 0.214 |
| | CRSsNP vs. CRSwNP | 0.010 |
| | Healthy vs. CRSwNP | 0.267 |
| **IL-5** | CRSsNP vs. Healthy | 0.573 |
| | CRSsNP vs. CRSwNP | 0.020 |
| | Healthy vs. CRSwNP | 0.136 |
| **IL-6** | CRSsNP vs. Healthy | 0.229 |
| | CRSsNP vs. CRSwNP | 0.021 |
| | Healthy vs. CRSwNP | 0.479 |
| **IL-8** | CRSsNP vs. Healthy | 0.867 |
| | CRSsNP vs. CRSwNP | 0.050 |
| | Healthy vs. CRSwNP | 0.184 |
| **IL-9** | CRSsNP vs. Healthy | 0.444 |
| | CRSsNP vs. CRSwNP | 0.015 |
| | Healthy vs. CRSwNP | 0.151 |
| **IL-10** | CRSsNP vs. Healthy | 0.544 |
| | CRSsNP vs. CRSwNP | 0.040 |
| | Healthy vs. CRSwNP | 0.305 |
| **IL-12p70** | CRSsNP vs. Healthy | 0.398 |
| | CRSsNP vs. CRSwNP | 0.037 |
| | Healthy vs. CRSwNP | 0.412 |
| **IL13** | CRSsNP vs. Healthy | 0.146 |
| | CRSsNP vs. CRSwNP | 0.023 |
| | Healthy vs. CRSwNP | 0.760 |
| **IL-37** | CRSsNP vs. Healthy | >0.999 |
| | CRSsNP vs. CRSwNP | 0.073 |

| | | |
|---|---|---|
| | Healthy vs. CRSwNP | 0.184 |
| **IP-10** | CRSsNP vs. Healthy | >0.999 |
| | CRSsNP vs. CRSwNP | >0.999 |
| | Healthy vs. CRSwNP | 0.933 |
| **MCP-1** | CRSsNP vs. Healthy | 0.496 |
| | CRSsNP vs. CRSwNP | 0.021 |
| | Healthy vs. CRSwNP | 0.181 |
| **MIP-1a** | CRSsNP vs. Healthy | 0.349 |
| | CRSsNP vs. CRSwNP | 0.030 |
| | Healthy vs. CRSwNP | 0.400 |
| **MIP-1ß** | CRSsNP vs. Healthy | 0.349 |
| | CRSsNP vs. CRSwNP | 0.030 |
| | Healthy vs. CRSwNP | 0.400 |
| **MMP-1** | CRSsNP vs. Healthy | 0.090 |
| | CRSsNP vs. CRSwNP | 0.009 |
| | Healthy vs. CRSwNP | 0.595 |
| **MMP-7** | CRSsNP vs. Healthy | 0.265 |
| | CRSsNP vs. CRSwNP | 0.018 |
| | Healthy vs. CRSwNP | 0.357 |
| **MMP-8** | CRSsNP vs. Healthy | 0.473 |
| | CRSsNP vs. CRSwNP | 0.010 |
| | Healthy vs. CRSwNP | 0.092 |
| **MPO** | CRSsNP vs. Healthy | 0.908 |
| | CRSsNP vs. CRSwNP | 0.034 |
| | Healthy vs. CRSwNP | 0.110 |
| **SAA** | CRSsNP vs. Healthy | 0.170 |
| | CRSsNP vs. CRSwNP | 0.010 |
| | Healthy vs. CRSwNP | 0.349 |
| **TNFa** | CRSsNP vs. Healthy | 0.481 |
| | CRSsNP vs. CRSwNP | 0.023 |
| | Healthy vs. CRSwNP | 0.206 |
| **TRAIL** | CRSsNP vs. Healthy | 0.447 |
| | CRSsNP vs. CRSwNP | 0.012 |
| | Healthy vs. CRSwNP | 0.122 |
| **TREM-1** | CRSsNP vs. Healthy | 0.135 |
| | CRSsNP vs. CRSwNP | 0.004 |
| | Healthy vs. CRSwNP | 0.220 |

8

**Supplementary Table 4: Kruskal-Wallis test comparing infectious cohorts, with Dunn's pairwise comparisons**

| Cytokine | Comparison Cohort | Adjusted P Value |
|---|---|---|
| **CRP** | Bacteria vs. Healthy | <0.001 |
| | Bacteria vs. Viral | 0.017 |
| | Healthy vs. Viral | 0.002 |
| **EGF** | Bacteria vs. Healthy | <0.001 |
| | Bacteria vs. Viral | 0.218 |
| | Healthy vs. Viral | 0.001 |
| **Eotaxin-1** | Bacteria vs. Healthy | <0.001 |
| | Bacteria vs. Viral | 0.101 |
| | Healthy vs. Viral | <0.001 |
| **Eotaxin-2** | Bacteria vs. Healthy | <0.001 |
| | Bacteria vs. Viral | 0.004 |
| | Healthy vs. Viral | 0.198 |
| **Eotaxin-3** | Bacteria vs. Healthy | <0.001 |
| | Bacteria vs. Viral | 0.011 |
| | Healthy vs. Viral | 0.084 |
| **Fractalkine** | Bacteria vs. Healthy | <0.001 |
| | Bacteria vs. Viral | 0.182 |
| | Healthy vs. Viral | 0.011 |
| **G-CSF** | Bacteria vs. Healthy | <0.001 |
| | Bacteria vs. Viral | 0.008 |
| | Healthy vs. Viral | 0.033 |
| **GM-CSF** | Bacteria vs. Healthy | <0.001 |
| | Bacteria vs. Viral | 0.018 |
| | Healthy vs. Viral | 0.109 |
| **GzmA** | Bacteria vs. Healthy | <0.001 |
| | Bacteria vs. Viral | 0.427 |
| | Healthy vs. Viral | <0.001 |
| **GzmB** | Bacteria vs. Healthy | <0.001 |
| | Bacteria vs. Viral | 0.025 |
| | Healthy vs. Viral | 0.008 |
| **IFNγ** | Bacteria vs. Healthy | 0.001 |
| | Bacteria vs. Viral | 0.015 |
| | Healthy vs. Viral | 0.378 |
| **IL-1RA** | Bacteria vs. Healthy | <0.001 |

| | | |
|---|---|---|
| | Bacteria vs. Viral | 0.106 |
| | Healthy vs. Viral | 0.040 |
| **IL-1a** | Bacteria vs. Healthy | <0.001 |
| | Bacteria vs. Viral | 0.031 |
| | Healthy vs. Viral | 0.017 |
| **IL-2** | Bacteria vs. Healthy | <0.001 |
| | Bacteria vs. Viral | 0.037 |
| | Healthy vs. Viral | 0.036 |
| **IL-3** | Bacteria vs. Healthy | <0.001 |
| | Bacteria vs. Viral | <0.001 |
| | Healthy vs. Viral | >0.999 |
| **IL-4** | Bacteria vs. Healthy | <0.001 |
| | Bacteria vs. Viral | 0.025 |
| | Healthy vs. Viral | 0.080 |
| **IL-5** | Bacteria vs. Healthy | <0.001 |
| | Bacteria vs. Viral | 0.017 |
| | Healthy vs. Viral | 0.046 |
| **IL-6** | Bacteria vs. Healthy | <0.001 |
| | Bacteria vs. Viral | 0.013 |
| | Healthy vs. Viral | 0.284 |
| **IL-8** | Bacteria vs. Healthy | <0.001 |
| | Bacteria vs. Viral | 0.019 |
| | Healthy vs. Viral | 0.016 |
| **IL-9** | Bacteria vs. Healthy | <0.0001 |
| | Bacteria vs. Viral | 0.010 |
| | Healthy vs. Viral | 0.039 |
| **IL-10** | Bacteria vs. Healthy | <0.001 |
| | Bacteria vs. Viral | 0.010 |
| | Healthy vs. Viral | 0.099 |
| **IL-12p70** | Bacteria vs. Healthy | <0.001 |
| | Bacteria vs. Viral | 0.017 |
| | Healthy vs. Viral | 0.315 |
| **IL13** | Bacteria vs. Healthy | 0.002 |
| | Bacteria vs. Viral | 0.015 |
| | Healthy vs. Viral | 0.506 |
| **IL-37** | Bacteria vs. Healthy | 0.004 |
| | Bacteria vs. Viral | >0.999 |

| | | |
|---|---|---|
| | Healthy vs. Viral | 0.001 |
| **IP-10** | Bacteria vs. Healthy | 0.040 |
| | Bacteria vs. Viral | >0.999 |
| | Healthy vs. Viral | <0.001 |
| **MCP-1** | Bacteria vs. Healthy | 0.006 |
| | Bacteria vs. Viral | >0.999 |
| | Healthy vs. Viral | <0.001 |
| **MIP-1a** | Bacteria vs. Healthy | <0.001 |
| | Bacteria vs. Viral | 0.099 |
| | Healthy vs. Viral | 0.005 |
| **MIP-1ß** | Bacteria vs. Healthy | <0.001 |
| | Bacteria vs. Viral | 0.099 |
| | Healthy vs. Viral | 0.013 |
| **MMP-1** | Bacteria vs. Healthy | <0.001 |
| | Bacteria vs. Viral | 0.012 |
| | Healthy vs. Viral | 0.027 |
| **MMP-7** | Bacteria vs. Healthy | <0.001 |
| | Bacteria vs. Viral | 0.069 |
| | Healthy vs. Viral | 0.002 |
| **MMP-8** | Bacteria vs. Healthy | <0.001 |
| | Bacteria vs. Viral | 0.011 |
| | Healthy vs. Viral | 0.114 |
| **MPO** | Bacteria vs. Healthy | <0.001 |
| | Bacteria vs. Viral | 0.010 |
| | Healthy vs. Viral | 0.162 |
| **SAA** | Bacteria vs. Healthy | 0.007 |
| | Bacteria vs. Viral | >0.999 |
| | Healthy vs. Viral | <0.001 |
| **TNFa** | Bacteria vs. Healthy | <0.001 |
| | Bacteria vs. Viral | 0.026 |
| | Healthy vs. Viral | 0.013 |
| **TRAIL** | Bacteria vs. Healthy | 0.028 |
| | Bacteria vs. Viral | >0.999 |
| | Healthy vs. Viral | <0.001 |
| **TREM-1** | Bacteria vs. Healthy | <0.001 |
| | Bacteria vs. Viral | 0.008 |
| | Healthy vs. Viral | 0.029 |

**Supplementary Table 5: Wilcoxon rank-sum test comparing type-2 and non-type 2 inflammation cohorts.**

| Cytokine | P value |
|---|---|
| **CRP** | 0.417 |
| **EGF** | 0.029 |
| **Eotaxin-1** | 0.463 |
| **Eotaxin-2** | 0.194 |
| **Eotaxin-3** | 0.247 |
| **Fractalkine** | 0.087 |
| **G-CSF** | 0.049 |
| **GM-CSF** | 0.217 |
| **GzmA** | 0.119 |
| **GzmB** | 0.583 |
| **IFNy** | 0.091 |
| **IL-1RA** | >0.999 |
| **IL-1a** | 0.042 |
| **IL-2** | 0.583 |
| **IL-3** | 0.068 |
| **IL-4** | 0.726 |
| **IL-5** | 0.436 |
| **IL-6** | 0.391 |
| **IL-8** | 0.139 |
| **IL-9** | 0.058 |
| **IL-10** | 0.241 |
| **IL-12p70** | 0.153 |
| **IL-13** | 0.808 |
| **IL-37** | 0.068 |
| **IP-10** | >0.999 |
| **MCP-1** | 0.019 |
| **MIP-1a** | 0.020 |
| **MIP-1ß** | 0.026 |
| **MMP-1** | 0.583 |
| **MMP-7** | 0.241 |
| **MMP-8** | 0.820 |
| **MPO** | 0.135 |
| **SAA** | 0.135 |

| | |
|---|---|
| **TNFa** | 0.104 |
| **TRAIL** | 0.715 |
| **TREM-1** | 0.199 |

14

**LIST OF ABBREVIATIONS**

ANOVA: Analysis of Variance

CCL11: C-C Motif Chemokine Ligand 11

CCL24: C-C Motif Chemokine Ligand 24

CCL26: C-C Motif Chemokine Ligand 26

COVID-19: Coronavirus Disease 2019

CRD: Chronic Respiratory Disease

CRS: Chronic Rhinosinusitis

CX3CL1: C-X3-C Motif Chemokine Ligand 1

EGF: Epidermal Growth Factor

G-CSF: Granulocyte Colony-Stimulating Factor

GM-CSF: Granulocyte Macrophage Colony-Stimulating Factor

GzmA: Granzyme A

GzmB: Granzyme B

IFN-α: Interferon-Alpha

IFNγ: Interferon-Gamma

IL-1RA: Interleukin-1 Receptor Antagonist

IL-2: Interleukin-2

IL-3: Interleukin-3

IL-4: Interleukin-4

IL-5: Interleukin-5

IL-6: Interleukin-6

IL-8: Interleukin-8

IL-9: Interleukin-9

IL-10: Interleukin-10

IL-12p70: Interleukin-12p70

IL-13: Interleukin-13

IL-22: Interleukin-22

IL-37: Interleukin-37

IP-10: Interferon Gamma-Induced Protein 10

MCP-1: Monocyte Chemoattractant Protein 1

MIP-1α: Macrophage Inflammatory Protein 1-Alpha

MIP-1β: Macrophage Inflammatory Protein 1-Beta

MMP-1: Matrix Metalloproteinase 1

MMP-7: Matrix Metalloproteinase 7

MMP-8: Matrix Metalloproteinase 8

MPO: Myeloperoxidase

SAA: Serum Amyloid A

SNOT-22: Sino-Nasal Outcome Test

SPA: Specialty Physician Associates

TNF-α: Tumor Necrosis Factor Alpha

TNFβ: Tumor Necrosis Factor Beta

TRAIL: TNF-Related Apoptosis Inducing Ligand

TREM-1: Triggering Receptor Expressed on Myeloid Cells 1

TREM-2: Triggering Receptor Expressed on Myeloid Cells 2

### DECLARATIONS

#### Ethics approval and consent to participate

Informed consent was obtained from patients.

#### Consent for publication

Not applicable

#### Availability of data and materials

All data generated or analysed during this study are included in this published article and its supplementary information files.

#### Competing Interests

All authors receive financial compensation from Diag-Nose Medical, which funded this research. As such, they may have a financial or professional interest in the results. This relationship is disclosed in the author affiliations. No additional competing interests are declared.

#### Funding

This work was funded by Diag-nose Medical.

#### Authors' contributions

TL conducted the formal analysis and contributed to conceptualisation, drafting, review and editing of the manuscript. ST contributed to the formal analysis and editing of the manuscript. AD contributed to the conceptualization and design of the study, and participated in manuscript review and editing. ER, DY, and BW contributed to the conceptualization and study design. All authors read and approved the final manuscript.

#### Acknowledgements

We thank L.Kaur and A.Yusim of Crux Biolabs for performing the experiments.
